# Mental Health Trends in India from 2020 to 2022: Association with Financial Stress, Food Insecurity, and COVID-19-related Illness Concerns

**DOI:** 10.1101/2024.03.17.24304396

**Authors:** Youqi Yang, Anqi Sun, Lauren Zimmermann, Bhramar Mukherjee

## Abstract

This study examines the impact of pandemic-related worries on mental health in the Indian general adult population from 2020 to 2022. Using data from the Global COVID-19 Trends and Impact Survey (N = 2,576,174 respondents aged *≥*18 years in India; an average weekly sample size of around 25,000), it explores the associations between worry variables (namely financial stress, food insecurity, and COVID-19-related health worries) and self-reported symptoms of depression and nervousness. The statistical analysis was conducted using complete cases only (N = 747,996). Our analysis used survey-weighted models, focusing on the three pandemic-related worries as the exposures, while also adjusting for various other covariates, including demographics and calendar time. The study finds significant associations between these worries and mental health outcomes, with financial stress being the most significant factor affecting both depression (adjusted odds ratio: 2.36, 95% confidence interval: [2.27, 2.46]) and nervousness (adjusted odds ratio: 1.91, 95% confidence interval: [1.81, 2.01]) during the first phase of the study period (June 27, 2020, to May 19, 2021). The fully adjusted models also identify additional factors related to mental health, including age, gender, residential status, geographical region, occupation, and education. Moreover, the research highlights that males and urban residents had higher odds ratios for self-reported mental health problems regarding the worry variables than females and rural residents, respectively. Furthermore, the study reveals a rise in the prevalence of self-reported depression and nervousness and their association with COVID-19-related health worries during the lethal second wave of the pandemic in May 2021 compared to the onset of the pandemic. This study shows that social media platforms like Facebook can deploy surveys to a large number of participants globally and can be useful tools in capturing mental health trends and uncovering associations during a public health crisis.

## 1 Introduction

The COVID-19 pandemic has presented an unprecedented challenge to not only the physical well-being but also the mental health of individuals (Vindegaard and Benros, 2020). Global surveys have consistently reported a surge in depression, anxiety, and stress symptoms linked to the COVID-19 pandemic (Cao et al., 2020; Gao et al., 2020; Qiu et al., 2020; Wang et al., 2020; González-Sanguino et al., 2020; Czeisler et al., 2020). A comprehensive review analyzing 41 studies from various countries, including China, Spain, Italy, Iran, the US, Turkey, Nepal, and Denmark, revealed relatively high occurrences of anxiety (ranging from 6.33% to 50.9%) and depression (ranging from 14.6% to 48.3%) (Xiong et al., 2020). This increase in mental health problems extends beyond individuals diagnosed with COVID-19, affecting the wider population as a result of psychosocial stressors like lifestyle disruptions, the fear of contracting the virus, and concerns over potential economic downturns (Moreno et al., 2020).

As the world’s second most populous nation, India’s handling of the COVID-19 pandemic is considered pivotal in contributing to the global response to the crisis (Mukherjee et al., 2021). Extant literature suggests that this crisis has been marked by a pronounced surge in the prevalence of adverse mental health among the Indian general population throughout the pandemic period (Dalal et al., 2020). A cross-sectional study conducted in April 2020 found that 40.5% of the respondents reported either anxiety or depression (Grover et al., 2020). This marked increase sharply contrasts with the pre-pandemic national depression rate of 2.7% estimated in the National Mental Health Survey of 2015 (Arvind et al., 2019). Several other studies have also emphasized the elevated levels of anxiety and depression experienced by the Indian population during the pandemic. These researchers explored a spectrum of factors associated with mental health outcomes, including age, gender, education, urban-rural residential status, states of residence, and occupations (Roy et al., 2020; Verma and Mishra, 2020; Gopal et al., 2020; Venugopal et al., 2022). Moreover, some studies concentrate on understanding symptoms of mental illness within specific subgroups in India, such as healthcare workers (Suryavanshi et al., 2020; Wilson et al., 2020b), college students (Verma, 2020), migrant laborers (Kumar et al., 2020), and pregnant women (Jungari, 2020).

In the early stages of the pandemic, the Indian government swiftly implemented a nationwide lockdown from March to May 2020, affecting the mobility of over 1.4 billion people. During the second wave in 2021, various states also enforced restrictions on their residents (Kar et al., 2021; Salvatore et al., 2022). Rehman et al. (2021) showed that being unprepared for lockdown was linked to an increase in mental health conditions. The stringent lockdown measures led to the closure of public services, commercial establishments, transportation, and educational facilities. This resulted in a dramatic spike in unemployment rates in India, escalating from 8 percent in March 2020 to 23 percent in May 2020, and further to 35 percent by June 2020, as reported by the Centre for Monitoring Indian Economy (Dev et al., 2020). The year 2020 also saw a significant downturn in the economy, marked by a 7% reduction in Gross Domestic Product (GDP), which was most pronounced in the first quarter with a steep decline of 24% (International Monetary Fund, 2023). These situations may lead to a decline in household income and a scarcity of necessary resources including food and other essentials. A large-scale study conducted in 2020 among 12 states in India found that approximately 80 percent of the households faced a decrease in their food consumption, while over 60 percent reported insufficient funds to cover a week’s worth of essential expenses (Kesar et al., 2021). Another longitudinal study conducted in Uttar Pradesh also reported a significant shift in food security status in 2020, with 62% of households transitioning from being food secure to food insecure (Nguyen et al., 2021). Financial hardship and food insecurity may also disproportionately impact vulnerable population segments, such as migrant laborers, street vendors, rural inhabitants, and women (Kesar et al., 2021; Mishra and Rampal, 2020).

Prior studies have revealed the associations between financial stress, food insecurity, and deteriorating mental health during the COVID-19 pandemic in other countries. Wilson et al. (2020a) found a significant association between heightened financial stress and increased depressive and anxiety symptoms in the United States. Parallel results were reported by Borrescio-Higa et al. (2022) from a cross-sectional survey conducted in Chile, Blix et al. (2021) from Norway, and Zajacova et al. (2020) from Canada. On the other hand, Fang et al. (2021) highlighted that food insecurity caused by the pandemic was linked to an increased risk of mental illness in the United States. The consistent findings were further affirmed in subsequent publications (Wolfson et al., 2021; Lauren et al., 2021; Fitzpatrick et al., 2021; Rahman et al., 2021). Despite existing research, there is still a notable gap in studies focusing on the general population in India. Lathabhavan (2022) highlighted financial stress as a key factor linked to depression and anxiety, although their research was confined to small business entrepreneurs. Similarly, Chatterji et al. (2021) reported comparable results, but the sample of their study was limited to a rural agrarian community in Maharashtra. Moreover, the fear of contracting COVID-19 has also emerged as another risk factor for adverse mental health symptoms. In a survey involving Indian college students, Lathabhavan and Vispute (2022) discovered an association between the fear of COVID-19-related illness and stress. Worries regarding the illness could significantly contribute to mental health and wellness in India, particularly given the country’s vulnerable healthcare system and inadequate investments during this unprecedented pandemic (Chetterje, 2020).

In this study, we used data from the Global COVID-19 Trends and Impact Survey (CTIS), conducted in collaboration between Meta/Facebook and the University of Maryland, spanning from 2020 to 2022 (Astley et al., 2021). With its broad coverage of COVID-19-related inquiries, which includes mental health assessments, this survey stands as a key resource for the investigation of mental health trajectories across the world during the pandemic. In the context of the United States, researchers have analyzed both county-level and individual-level data to assess the factors associated with anxiety, depression, and isolation during the pandemic (Lupton-Smith et al., 2022; Ettman et al., 2023; Kobayashi et al., 2023). Additionally, Kush et al. (2022) observed a higher prevalence of anxiety symptoms among teachers compared to other professions. Expanding the lens globally, Botha et al. (2023) conducted a study on the trajectories of depression and anxiety across five Australian states from 2020 to 2021. Their research highlighted the mediating roles of financial stress and COVID-19-related illness concerns in the effect of the lockdown on mental health conditions. However, the majority of ongoing studies using the CTIS are primarily focused on the United States, whereas little or no attention has been given to low- or middle-income countries (LMICs) like India. Furthermore, while prior research in India has promptly assessed mental health status during the pandemic’s initial phase (Grover et al., 2020; Roy et al., 2020), there is a dearth of knowledge regarding mental health trends throughout subsequent periods of the pandemic, which is crucial for assessing the lingering effects of COVID-19.

The primary objective of this research is to investigate the mental health dynamics among Indian adults over the period 2020-2022 during the pandemic. We consider depression and nervousness as the primary outcomes and explore their possible associations with pandemic-related worries, including financial stress, food insecurity, and COVID-19-related illness concerns, after adjusting for demographic variables and calendar time.

The structure of this paper is outlined as follows: Section 2 details the study design, the measures of the variables, and the statistical methods employed in our analysis. Section 3 presents the exploratory and analytical results. Finally, Section 4 delves into a discussion of the reported findings and limitations of the current study.

## 2 Materials and methods

### 2.1 Study design

In our study, we used non-aggregated data sourced from the Global COVID-19 Trends and Impact Survey (CTIS) (Astley et al., 2021). Daily, it invited a stratified random sample of adults from the Facebook Active User Base (FAUB) to complete a cross-sectional questionnaire. The questionnaire covered various aspects of the COVID-19 pandemic, including mental health, pandemic-related worries, demographics, and more. To ensure the survey respondents accurately represented the general adult population, Facebook implemented a two-fold weighting strategy (Barkay et al., 2020; The Delphi Group at Carnegie Mellon University and University of Maryland Social Data Science Center, in partnership with Meta, 2022). First, non-response weights were calculated using an inverse propensity score weighting method. This adjustment was made to mitigate potential non-response bias, thereby enhancing the representativeness of the survey participants within the FAUB. Second, post-stratification weights were applied to align the survey data with the United Nations (UN) Population Division’s 2019 World Population Projection in terms of age and gender. This step was to address the non-coverage bias arising from inactive Facebook users or those without internet access. For our research, we accessed individual response data and the corresponding weights through a Data User Agreement with the University of Maryland from its data repository (University of Maryland, 2022).

The CTIS initiated inquiries into mental health outcomes on April 23, 2020, and expanded to cover financial stress, food insecurity, and COVID-19-related health concerns starting June 27, 2020. Our study used responses from June 27, 2020 (the first date when the relevant data became available), up to the last data collection day on June 25, 2022 (N = 2,576,174 adult respondents in India; average sample size around 25,000/week). A major alteration in survey structure occurred on May 20, 2021, involving the randomization of respondents into two different question modules instead of responding to all questions. Specifically, questions pertaining to mental health, financial stress, and food insecurity were included in only one of these two modules, and inquiries about COVID-19 health concerns were phased out from the survey. This structural shift resulted in discontinuities in several survey estimates (University of Maryland, 2022), prompting us to divide our analysis into two distinct periods: June 27, 2020, to May 19, 2021 (referred to as Period 1; N = 1,532,776), and May 20, 2021, to June 25, 2022 (referred to as Period 2; N = 1,043,398). Only complete cases from both Period 1 (N = 595,229) and Period 2 (N = 152,767) were used for modeling (Figure S1).

### 2.2 Measures

#### Outcomes

We investigated two commonly studied mental health outcomes: depression and nervousness. Self-reported depression was assessed with the question, “During the last 7 days, how often did you feel so depressed that nothing could cheer you up?” For self-reported nervousness, participants were asked, “During the last 7 days, how often did you feel so nervous that nothing could calm you down?” Responses were recorded on a 5-point scale: 1 = “all the time”, 2 = “most of the time”, 3 = “some of the time”, 4 = “a little of the time”, 5 = “none of the time”. These questions were adapted from the Center for Epidemiological Studies Depression Scale (CES-D) and the Generalized Anxiety Disorder-7 (GAD-7), which are widely acknowledged for screening depression and nervousness, respectively (Radloff, 1977; Spitzer et al., 2006). These short-form, single-item adaptations of the aforementioned scales reflect self-reported symptoms of depression and nervousness and do not reflect clinical diagnoses of related psychiatric health conditions (e.g., major depressive disorder and general anxiety disorder). The phrasing of the survey questions and the initial participant responses are detailed in Table S1. The original responses to the depression and nervousness inquiries were moderately correlated (Kendall rank correlation coefficient = 0.59 and 0.58 in Period 1 and Period 2, respectively). For the analysis, we dichotomized the responses into binary indicators: 0 for “some, a little, or none of the time”, and 1 for “all or most of the time”, aligning with prior research (Kush et al., 2022; Ettman et al., 2023; Botha et al., 2023).

#### Primary exposures

The primary exposures were subjective worries about the pandemic, assessed through three aspects: financial stress, food insecurity, and COVID-19-related illness concerns (Table S1). Financial stress was assessed using the question, “How worried are you about your household’s finances in the next month?”. For food insecurity, participants were asked, “How worried are you about having enough to eat in the next week?” In terms of COVID-19-related illness concerns, the question posed was, “How worried are you that you or someone in your immediate family might become seriously ill from coronavirus (COVID-19)?” Responses were recorded on a 4-point scale: 1 = “very worried”, 2 = “somewhat worried”, 3 = “not too worried”, 4 = “not worried at all”. For the analysis, we dichotomized the responses into binary indicators: 0 for “somewhat worried, not too worried, or not worried at all”, and 1 for “very worried”.

#### Other covariates

Demographic information often used as covariates was also collected from the respondents (Table S1). Gender was self-reported and categorized into “male”, “female”, “other”, and “prefer not to answer”. In our analysis, we considered gender as a binary variable, excluding those who reported “other” (N = 1,593; 0.01%) or preferred not to answer (N = 12,534; 0.05%). Age was categorized into groups: 18-24, 25-34, 35-44, 45-54, 55-64, 65-74, and 75+ years old. Due to the limited sample size, we combined 65-74 and 75+ years old into a group of 65+ years old. Regarding educational attainment, participants were initially asked to report the number of years of education completed via an open-ended question in Period 1. In Period 2, this was changed to a multiple-choice question asking for the highest level of education completed. To mitigate potential measurement errors, we only included non-negative responses below a reasonable threshold (50 years) in Period 1 for analysis. For consistency, we dichotomized education responses in the two periods into binary indicators: 0 for less than a high school degree (less than 12 years) and 1 for a high school degree or more (12 years or more). The residential status was converted from a categorical response to a binary indicator: 0 denoting rural settings (village or rural), and 1 denoting urban environments (city or town). Respondents reported their Indian state or territory of residence, which we grouped into six regions based on the divisions by the zonal councils. Occupations were analyzed as a categorical variable, including agriculture, construction, education, health, tourism, transportation, and others.

### 2.3 Statistical methods

For the exploratory analysis, we utilized the full sample for both periods (Figure S1). First, we calculated both unweighted and weighted summary statistics for demographic factors, worries about the pandemic, and mental health outcomes. We used weights provided by Facebook to account for non-response and non-coverage bias, as mentioned in the previous section. In addition, we provided demographic proportions among FAUB, estimated by the Facebook advertising tools (Facebook, 2023), and those of the general Indian adult population, sourced from the latest Census (2011) or National Sample Survey (Government of India, 2011). Second, to evaluate the unadjusted correlations among worry variables without weights, we computed the phi coefficients, given that these variables were categorized as binary. Third, we generated time-series plots illustrating the trajectories of depression and nervousness over time. These plots were constructed using weekly weighted proportions after computing the 7-day rolling averages. To complement these visualizations, we also presented time-series plots depicting weekly new confirmed COVID-19 cases and weekly new COVID-19-related deaths in India, sourced from the Johns Hopkins Coronavirus Resource Center (Dong et al., 2020).

For the statistical analysis, we fitted separate survey-weighted logistic regression models for depression and nervousness as outcomes within each period, using the complete cases only. First, we constructed unadjusted models, each incorporating a single covariate from pandemic-related worries (financial stress, food insecurity, and COVID-19-related illness concerns), demographics (gender, age, education, urban-rural residential status, region, and occupation), and calendar time (categorized by month and year). Second, we fit partially adjusted models with a single exposure (namely using one worry variable at a time) and all other covariates (demographics and time). Third, we formulated fully adjusted models integrating all three worry variables simultaneously and adjusting for the covariates. Fourth, we added the interaction term between gender and each worry exposure separately within the fully adjusted models, aiming to examine whether the association between pandemic-related worries and mental health outcomes varies by gender. Fifth, we explored whether the associations between exposures and mental health symptoms were modified by reported urban vs. rural residential status, by adding the interaction term between residential status and each of the worry exposures separately to the fully adjusted model. Sixth, we assessed the interaction between time and pandemic-related worries on psychological distress, by introducing time interactions with each of the three primary exposures separately, alongside the fully adjusted models. We employed robust sandwich estimators for variance and conducted the Wald test to investigate statistical significance. The analysis was performed using the survey R package (Lumley, 2004) in R version 4.3.2.

## 3 Results

### 3.1 Descriptive statistics

Table 1 presents a summary of both unweighted and weighted statistics for the variables within the full sample (N = 2,576,174 respondents aged *≥*18 years in India). Regarding mental health outcomes, the weighted proportions of respondents experiencing feelings of depression and nervousness were 8% and 6%, respectively, in both periods. In terms of subjective worries, in Period 1, 21% of the respondents post-weighted reported financial stress, 8% reported food insecurity, and 24% reported COVID-19-related illness concerns. Period 2 showed similar weighted statistics (17% for financial stress; 8% for food insecurity), except that COVID-19-related illness concerns were discontinued from the survey at this time. Notably, the demographics of the CTIS respondents, even after applying weights (considering age and gender), did not align accurately with the general Indian adult population. Specifically, the weighted sample had a larger proportion of males (66% in Period 1 and 57% in Period 2, as opposed to 52% in the census), people aged between 25 and 34 years (37% in Period 1 and 31% in Period 2, compared to 26% in the census), people having education levels of at least high school (83% in Period 1 and 93% in Period 2, as opposed to 14% in the census), and people residing in urban areas (75% in Period 1 and 75% in Period 2, compared to 31% in the census).

**Table 1:** Summary of unweighted and weighted statistics for demographics, worries about the pandemic, and mental health in the full sample (N = 2,576,174). Proportions for each category were calculated for categorical and binary variables, along with a count of missing data for each variable. Abbreviations: FAUB, Facebook Active User Base; HS, high school. a: Respondents (N = 386,356) randomized to another module were included in the missing value count for worry variables and mental health outcomes. b: Data were collected using Facebook advertising tools, providing information on gender, age, education, and region. c: Data were sourced from the latest census or National Sample Surveys, including information on gender (recorded as biological sex), age, education, residential status, and region. d: “Other” or “preferred not to answer” responses were treated as missing data. The survey participants provided self-reported gender information, whereas the census data reflects biological sex.

|  | Period 1 |  | Period 2<sup>a</sup> |  | FAUB<sup>b</sup><br>(2022) | Census<sup>c</sup> |
| --- | --- | --- | --- | --- | --- | --- |
|  | Jun 2020 - May 2021 |  | May 2021 - Jun 2022 |  |  |  |
|  | (N = 1,532,776) |  | (N = 1,043,398) |  |  |  |
| Variables | Unweighted | Weighted | Unweighted | Weighted |  |  |
| Demographics |  |  |  |  |  |  |
| Gender<sup>d</sup> |  |  |  |  |  |  |
| Female | 17% | 34% | 17% | 43% | 24% | 48% |
| Male | 83% | 66% | 83% | 57% | 76% | 52% |
| (Missing) | 597,175 |  | 334,133 |  |  |  |
| Age |  |  |  |  |  |  |
| 18-24 | 22% | 21% | 20% | 17% | 37% | 14% |
| 25-34 | 43% | 37% | 38% | 31% | 36% | 26% |
| 35-44 | 19% | 16% | 21% | 15% | 16% | 21% |
| 45-54 | 8% | 14% | 11% | 18% | 6% | 17% |
| 55-64 | 4% | 8% | 6% | 11% | 3% | 12% |
| 65+ | 2% | 5% | 5% | 8% | 2% | 10% |
| (Missing) | 578,727 |  | 328,421 |  |  |  |
| Education |  |  |  |  |  |  |
| HS Or More | 83% | 83% | 92% | 93% | 67% | 14% |
| Less than HS | 17% | 17% | 8% | 7% | 33% | 86% |
| (Missing) | 660,043 |  | 337,634 |  |  |  |
| Residential Status |  |  |  |  |  |  |
| Rural | 26% | 25% | 29% | 25% |  | 69% |
| Urban | 74% | 75% | 71% | 75% |  | 31% |
| (Missing) | 615,717 |  | 340,369 |  |  |  |
| Region |  |  |  |  |  |  |
| Central | 14% | 21% | 16% | 24% | 20% | 27% |
| East | 18% | 22% | 19% | 22% | 17% | 23% |

Table 1 continued from previous page
|  | Period 1 |  | Period 2 |  |  |  |
| --- | --- | --- | --- | --- | --- | --- |
|  | Jun 2020 - May 2021 |  | May 2021 - Jun 2022 |  |  |  |
|  | (N = 1,532,776) |  | (N = 1,043,398) |  |  |  |
| North | 21% | 15% | 21% | 14% | 19% | 13% |
| Northeast | 5% | 4% | 5% | 4% | 4% | 4% |
| South | 29% | 23% | 25% | 22% | 22% | 20% |
| West | 13% | 14% | 13% | 14% | 18% | 14% |
| (Missing) | 65,407 |  | 0 |  |  |  |
| Occupation |  |  |  |  |  |  |
| Agriculture | 8% | 9% | 12% | 12% |  |  |
| Construction | 5% | 4% | 5% | 4% |  |  |
| Education | 13% | 17% | 11% | 16% |  |  |
| Health | 8% | 8% | 7% | 8% |  |  |
| Tourism | 63% | 59% | 61% | 57% |  |  |
| Transportation | 2% | 1% | 1% | 1% |  |  |
| Others | 3% | 2% | 3% | 3% |  |  |
| (Missing) | 809,677 |  | 747,561 |  |  |  |
| Worries About The Pandemic |  |  |  |  |  |  |
| Financial Stress |  |  |  |  |  |  |
| Not Worried | 77% | 79% | 80% | 83% |  |  |
| Worried | 23% | 21% | 20% | 17% |  |  |
| (Missing) | 504,130 |  | 694,804 |  |  |  |
| Food Insecurity |  |  |  |  |  |  |
| Not Worried | 92% | 92% | 92% | 92% |  |  |
| Worried | 8% | 8% | 8% | 8% |  |  |
| (Missing) | 505,849 |  | 694,892 |  |  |  |
| COVID-19-related Illness Concern |  |  |  |  |  |  |
| Not Worried | 75% | 76% |  |  |  |  |
| Worried | 25% | 24% |  |  |  |  |
| (Missing) | 510,338 |  |  |  |  |  |
| Mental Health |  |  |  |  |  |  |
| Depression |  |  |  |  |  |  |
| Yes | 8% | 8% | 9% | 8% |  |  |
| No | 92% | 92% | 91% | 92% |  |  |
| (Missing) | 450,298 |  | 681,320 |  |  |  |
| Nervousness |  |  |  |  |  |  |
| Yes | 5% | 6% | 7% | 6% |  |  |
| No | 95% | 94% | 93% | 94% |  |  |

**Table 1 continued from previous page**
|  | **Period 1** | **Period 2** |
| --- | --- | --- |
|  | **Jun 2020 - May 2021** | **May 2021 - Jun 2022** |
|  | **(N = 1,532,776)** | **(N = 1,043,398)** |
| (Missing) | 450,656 | 680,232 |

We examined the unadjusted correlations among primary exposures without weights, using the full sample (N = 2,576,174). We found that binary indicators representing worry-related variables, specifically financial stress, and food insecurity, showed a moderate correlation with a phi coefficient of 0.49 without weighting. In Period 1, the correlation between financial stress and COVID-19-related illness concerns was quantified by a phi coefficient of 0.26, while the correlation between food insecurity and illness concerns was slightly higher at 0.29.

Figure 1 illustrates the weekly weighted prevalence of depression and nervousness from June 27, 2020, to June 25, 2022, using the full sample (N = 2,576,174). The vertical dashed line marks the time of the major change in the survey structure. Notably, there were three peaks in both the weekly new cases and deaths during the study period, occurring approximately in September 2020, May 2021, and January 2022. During the second wave around May 2021, the prevalence of both depression (10.11%) and nervousness (7.61%) symptoms also showed peaks, suggesting a simultaneous surge in mental health crises during the peak of the second lethal COVID-19 wave. However, this pattern was not as prominent during the first and third waves. When comparing the weighted proportions of individuals experiencing depression and nervousness at the start and end of the study period, the values in 2022 (8.29% for depression and 6.17% for nervousness) were higher than those observed in 2020 (7.24% for depression and 4.77% for nervousness). This underscores the enduring impact of the pandemic-related concerns on mental health within the Indian population.

**Figure 1:**
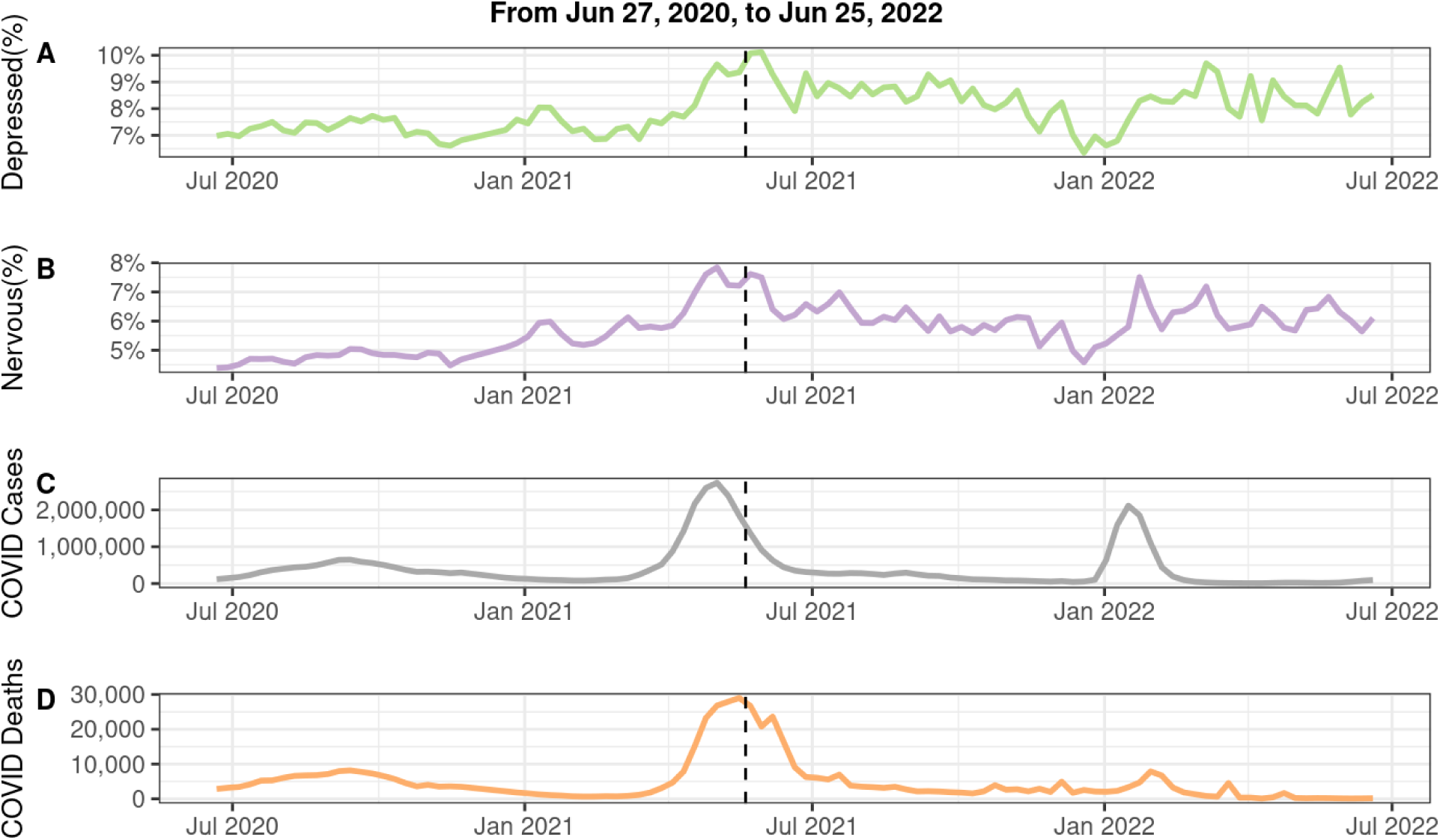
Weekly proportions of depression and nervousness, post-weighting, alongside weekly total of new COVID-19 cases and COVID-19-related deaths, from June 27, 2020, to June 25, 2022 (N = 2,576,174). (A) Weekly averages of weighted depression proportions; (B) weekly averages of weighted nervousness proportions; (C) weekly total of new COVID-19 confirmed cases; (D) weekly total of new COVID-19-related deaths. A 7-day smoothing method was employed before calculating the weighted averages. The significant alteration in the survey structure on May 20, 2021, is indicated by the vertical dashed line. The information on new COVID-19 confirmed cases and related deaths was obtained from the Johns Hopkins Coronavirus Resource Center.

Our gender-stratified analysis revealed notable distinctions in the weighted proportions. Regarding self-reported mental health problems, females showed a higher prevalence of depression than males in both Period 1 (10% vs. 6%) and Period 2 (9% vs. 8%), as well as increased nervousness in Period 1 (7% vs. 5%). Regarding the worry variables, males reported greater concerns about their financial situation (23% vs. 18% in Period 1; 19% vs. 14% in Period 2) and food status (9% vs. 6% in Period 1; 9% vs. 5% in Period 2) compared to females.

Likewise, we examined urban-rural differences in weighted proportions. In Period 1, the incidence of self-reported depression and nervousness among rural residents mirrored that of urban dwellers (8% vs. 8% for depression and 5% vs. 5% for nervousness), whereas Period 2 saw a higher occurrence in rural areas (9% vs. 8% for depression and 7% vs. 5% for nervousness). Concerning pandemic-related worries, rural individuals consistently reported greater levels of financial stress (28% vs. 19% in Period 1; 25% vs. 15% in Period 2) and food insecurity (12% vs. 7% in Period 1; 13% vs. 6% in Period 2) compared to urban residents.

### 3.2 Associations between worry variables and mental health outcomes

Table 2 displays the odds ratios for worry variables (considered as exposures) obtained from weighted logistic regression analyses. The analysis was based on complete cases from two distinct periods: Period 1 (N = 595,229) and Period 2 (N = 152,767). Here, we first report results from the fully adjusted models that concurrently incorporated all worry variables and other covariates, namely demographic factors and calendar time. The results from the fully adjusted models in Period 1 indicate significant associations between all three subjective worries (financial stress, food insecurity, and COVID-19-related illness worries) and mental health problems (both depression and nervousness). Notably, financial stress emerged as the most influential factor among the three worries. In particular, the odds of experiencing depression and nervousness were 2.36 (95% confidence interval, CI: [2.27, 2.46]) and 1.91 (95% CI: [1.81, 2.01]) times higher for those concerned about their financial situation compared to those without such worries, after adjusting for other variables, respectively. Individuals reporting food insecurity had higher adjusted odds for depression (odds ratio: 1.45, 95% CI: [1.38, 1.53]) and nervousness (odds ratio: 1.56, 95% CI: [1.47, 1.66]) compared to those without this concern. Similarly, the adjusted odds ratios regarding COVID-19-related illness worries were 1.65 (95% CI: [1.59, 1.71]) for depression and 1.81 (95% CI: [1.73, 1.90]) for nervousness.

**Table 2:**
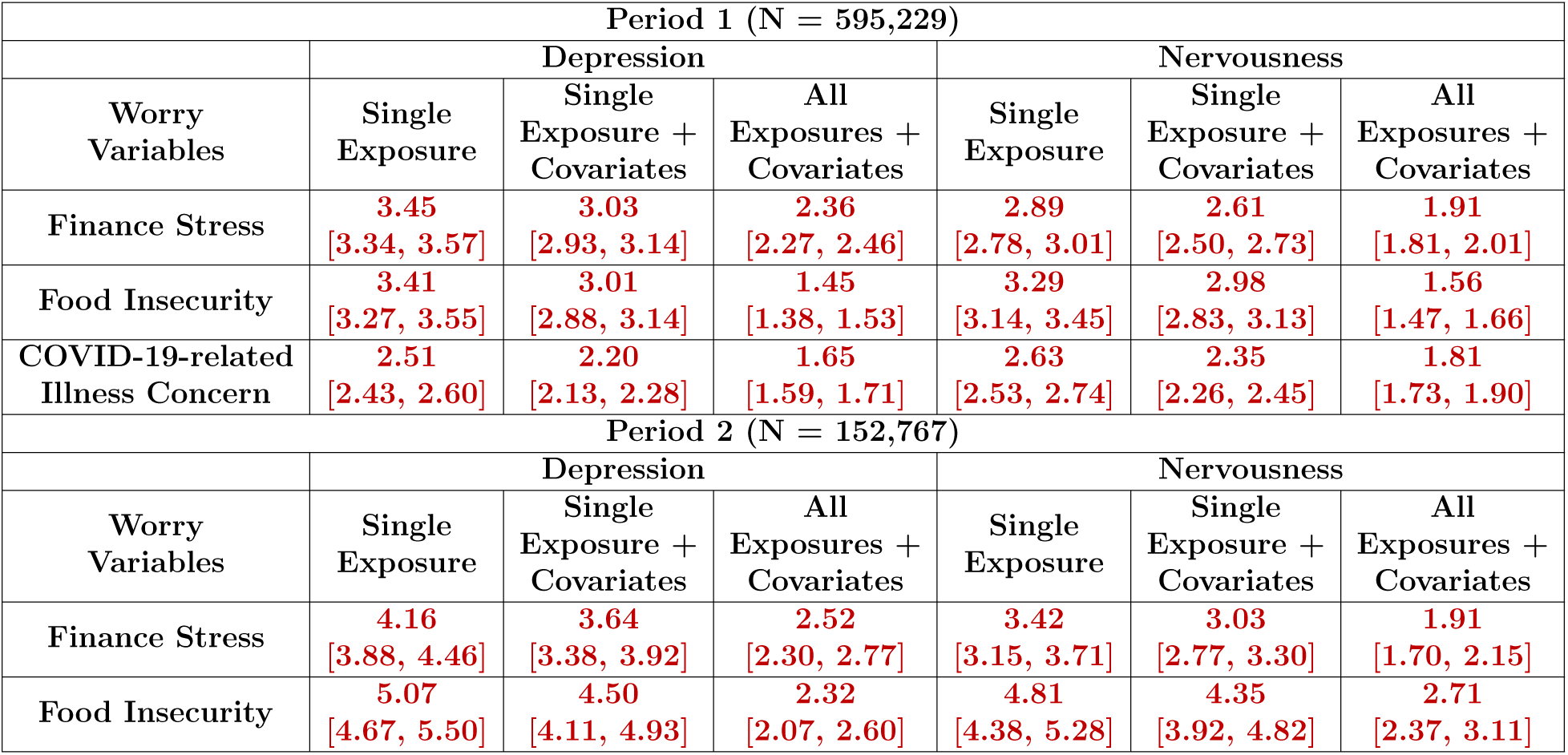
Fully adjusted, partially adjusted, and unadjusted model results for depression and nervousness in Period 1 (N = 595,229) and Period 2 (N = 152,767). The unadjusted models include one single exposure, selected from finance stress, food insecurity, and COVID-19-related illness concerns (not available in Period 2). The partially adjusted models include one single exposure and other covariates (demographics and time). The fully adjusted models include all of the three worry variables and other covariates (demographics and time) together. All model types integrate survey weights within logistic regression. Odds ratios are displayed as estimates with corresponding 95% Wald confidence intervals in the form of estimates [95% confidence interval], using a robust sandwich estimator for variance calculation. Significant results from Wald tests (significance level: 0.05) are highlighted in red and bold.

As depicted in the second part of Table 2, both financial stress and food insecurity had significant associations with mental health problems in the fully adjusted models in Period 2, mirroring findings from Period 1. Particularly, after accounting for all other variables, individuals experiencing financial stress had higher odds of feeling depressed (odds ratio: 2.52, 95% CI: [2.30, 2.77]) and nervous (odds ratio: 1.91, 95% CI: [1.70, 2.15]) compared to those without financial stress. Likewise, individuals reporting concerns about food insecurity exhibited significantly elevated adjusted odds of experiencing depression (odds ratio: 2.32, 95% CI: [2.07, 2.60]) and nervousness (odds ratio: 2.71, 95% CI: [2.31, 3.11]) compared to those without this concern.

In addition to the fully adjusted models, Table 2 presents outcomes from two other model types for each worry variable: unadjusted models, which only include a single exposure, and partially adjusted models, which include one exposure at a time together with demographic factors and calendar time. The comparisons across the three models for both periods highlight differences in the results. Notably, the odds ratios for all worry variables, consistently significant across models, were higher in the unadjusted models for both depression and anxiety. These values diminished in the partially adjusted models and were further reduced in the fully adjusted models. Taking financial stress as an example, in Period 1, individuals reporting financial stress had a 3.45 (95% CI: [3.34, 3.57]) times higher odds of experiencing depression compared to those without such worries, as indicated by the unadjusted model. This odds ratio decreased to 3.03 (95% CI: [2.93, 3.14]) with adjustments for demographics and calendar time, and further dropped to 2.36 (95% CI: [2.27, 2.46]) with additional adjustments for food insecurity and concerns about COVID-19-related illness. The ranking of the worry variables by their odds ratios also varied depending on whether adjustments were made, even within the same period and for the same outcome. For instance, in the first period, food insecurity showed the highest odds ratios for experiencing nervousness in both unadjusted (odds ratio: 3.29, 95% CI: [3.14, 3.45]) and partially adjusted models (odds ratio: 2.98, 95% CI: [2.83, 3.13]). Yet, in the fully adjusted models, the odds ratio for food insecurity (odds ratio: 1.56, 95% CI: [1.47, 1.66]) was lower than that for financial stress (odds ratio: 1.91, 95% CI: [1.81, 2.01]).

### 3.3 Associations between other demographics covariates and mental health outcomes

Table 3 and Table 4 present odds ratios of other covariates derived from weighted logistic regression models for both unadjusted models, featuring a single covariate, and fully adjusted versions that incorporate a complete set of relevant covariates, including the collective inclusion of all three worry variables. In Period 1, the analysis of fully adjusted models, as presented in Table 3, revealed significant relationships between demographic factors—such as gender, age, urban-rural residential status, geographical region, and occupation—and the prevalence of depression or nervousness. Key findings indicate that factors associated with poorer mental health outcomes include being female, being in the young adult age group, residing in urban areas, hailing from the southern regions, and engaging in occupations outside the agricultural sector. Notably, after controlling for other variables, individuals employed in the education sector had 1.23 (95% CI: [1.14, 1.34]) and 1.30 (95% CI: [1.18, 1.43]) times higher odds of reporting depression and nervousness, respectively, compared to those in agriculture. Similarly, those working in the tourism industry faced 1.55 (95% CI: [1.36, 1.77]) and 1.67 (95% CI: [1.24, 1.71]) times higher odds of experiencing depression and nervousness, respectively, in comparison to agricultural workers.

**Table 3:** Fully adjusted and unadjusted model results for depression and nervousness in Period 1 (N = 595,229). The unadjusted models include one single covariate, selected from pandemic-related worries, demographics, and time. In contrast, the fully adjusted models include all of the three worry variables and other covariates (demographics and time) together. Both model types integrate survey weights within logistic regression. Odds ratios are displayed as estimates with corresponding 95% Wald confidence intervals in the form of estimates [95% confidence interval], using a robust sandwich estimator for variance calculation. Significant results from Wald tests (significance level: 0.05) are highlighted in red and bold. Abbreviations: Ref, reference group; HS, high school. a: The June 2020 data were consolidated with the July 2021 dataset due to limitations in the sample size.

|  | Depression |  | Nervousness |  |
| --- | --- | --- | --- | --- |
|  | Unadjusted | Adjusted | Unadjusted | Adjusted |
| **Gender**<br>(Ref: Male) |  |  |  |  |
| Female | **1.66**<br>[1.60, 1.72] | **1.52**<br>[1.46, 1.59] | **1.55**<br>[1.48, 1.62] | **1.35**<br>[1.28, 1.42] |
| **Age**<br>(Ref: 18-24) |  |  |  |  |
| 25-34 | **0.70**<br>[0.67, 0.73] | **0.72**<br>[0.69, 0.75] | **0.75**<br>[0.71, 0.79] | **0.76**<br>[0.72, 0.80] |
| 35-44 | **0.43**<br>[0.40, 0.45] | **0.47**<br>[0.45, 0.50] | **0.52**<br>[0.49, 0.56] | **0.57**<br>[0.53, 0.60] |
| 45-54 | **0.24**<br>[0.22, 0.26] | **0.30**<br>[0.27, 0.32] | **0.38**<br>[0.35, 0.41] | **0.44**<br>[0.40, 0.48] |
| 55-64 | **0.16**<br>[0.13, 0.18] | **0.23**<br>[0.19, 0.26] | **0.31**<br>[0.27, 0.37] | **0.41**<br>[0.35, 0.48] |
| 65+ | **0.12**<br>[0.09, 0.16] | **0.19**<br>[0.14, 0.24] | **0.18**<br>[0.14, 0.24] | **0.25**<br>[0.19, 0.33] |
| **Education**<br>(Ref: HS or More) |  |  |  |  |
| Less than HS | **1.35**<br>[1.30, 1.40] | 1.01<br>[0.97, 1.06] | **1.30**<br>[1.25, 1.37] | 1.05<br>[1.00, 1.10] |
| **Residential Status**<br>(Ref: Urban) |  |  |  |  |
| Rural | **0.95**<br>[0.91, 0.98] | **0.82**<br>[0.79, 0.85] | 0.96<br>[0.92, 1.00] | **0.89**<br>[0.84, 0.93] |
| **Region**<br>(Ref: South) |  |  |  |  |
| Central | **1.08**<br>[1.03, 1.14] | **0.89**<br>[0.84, 0.94] | **0.93**<br>[0.87, 1.00] | **0.76**<br>[0.71, 0.82] |
| East | **1.34**<br>[1.28, 1.40] | **1.12**<br>[1.06, 1.17] | **1.09**<br>[1.03, 1.15] | **0.91**<br>[0.85, 0.96] |

Table 3 continued from previous page
|  | Depression |  | Nervousness |  |
| --- | --- | --- | --- | --- |
| North | 0.98<br>[0.94, 1.03] | **0.92**<br>**[0.88, 0.97]** | **0.81**<br>**[0.76, 0.86]** | **0.75**<br>**[0.71, 0.80]** |
| Northeast | 1.01<br>[0.93, 1.11] | **0.89**<br>**[0.82, 0.98]** | **0.89**<br>**[0.80, 0.99]** | **0.79**<br>**[0.71, 0.89]** |
| West | 0.97<br>[0.93, 1.02] | **0.91**<br>**[0.86, 0.96]** | 0.97<br>[0.91, 1.02] | **0.89**<br>**[0.84, 0.95]** |
| **Occupation**<br>(Ref: Agriculture) |  |  |  |  |
| Construction | **1.16**<br>**[1.05, 1.28]** | **1.17**<br>**[1.05, 1.29]** | **1.44**<br>**[1.32, 1.58]** | **1.29**<br>**[1.14, 1.45]** |
| Education | **1.43**<br>**[1.33, 1.54]** | **1.23**<br>**[1.14, 1.34]** | **1.31**<br>**[1.16, 1.46]** | **1.30**<br>**[1.18, 1.43]** |
| Health | 1.05<br>[0.97, 1.14] | 1.08<br>[0.99, 1.19] | **1.26**<br>**[1.13, 1.39]** | **1.30**<br>**[1.16, 1.45]** |
| Tourism | **1.76**<br>**[1.55, 1.99]** | **1.55**<br>**[1.36, 1.77]** | **1.31**<br>**[1.21, 1.41]** | **1.46**<br>**[1.24, 1.71]** |
| Transportation | **1.32**<br>**[1.18, 1.47]** | **1.20**<br>**[1.07, 1.35]** | **1.63**<br>**[1.41, 1.90]** | **1.21**<br>**[1.05, 1.38]** |
| Others | **1.26**<br>**[1.18, 1.34]** | **1.23**<br>**[1.15, 1.32]** | **1.33**<br>**[1.17, 1.52]** | **1.28**<br>**[1.17, 1.39]** |
| **Time**<br>(Ref: Jul-20)<sup>a</sup> |  |  |  |  |
| Aug-20 | 1.02<br>[0.97, 1.06] | **1.06**<br>**[1.01, 1.11]** | 1.02<br>[0.97, 1.08] | **1.07**<br>**[1.01, 1.13]** |
| Sep-20 | 1.04<br>[0.99, 1.09] | **1.12**<br>**[1.06, 1.17]** | **1.09**<br>**[1.03, 1.16]** | **1.16**<br>**[1.09, 1.23]** |
| Oct-20 | 1.02<br>[0.97, 1.08] | **1.14**<br>**[1.08, 1.20]** | 1.03<br>[0.96, 1.10] | **1.13**<br>**[1.05, 1.21]** |
| Nov-20 | **0.93**<br>**[0.87, 1.00]** | 1.04<br>[0.97, 1.12] | 0.98<br>[0.90, 1.06] | 1.08<br>[0.99, 1.18] |
| Dec-20 | 0.97<br>[0.90, 1.06] | 1.06<br>[0.98, 1.15] | **1.12**<br>**[1.02, 1.23]** | **1.20**<br>**[1.09, 1.32]** |
| Jan-21 | 1.03<br>[0.95, 1.11] | **1.17**<br>**[1.09, 1.27]** | **1.19**<br>**[1.09, 1.31]** | **1.33**<br>**[1.21, 1.46]** |
| Feb-21 | 0.95<br>[0.88, 1.03] | **1.12**<br>**[1.04, 1.21]** | 1.08<br>[0.98, 1.19] | **1.24**<br>**[1.12, 1.36]** |
| Mar-21 | 1.04<br>[0.97, 1.11] | **1.19**<br>**[1.11, 1.27]** | **1.28**<br>**[1.18, 1.39]** | **1.42**<br>**[1.31, 1.55]** |
| Apr-21 | **1.13**<br>**[1.07, 1.20]** | **1.16**<br>**[1.10, 1.24]** | **1.53**<br>**[1.43, 1.64]** | **1.54**<br>**[1.43, 1.64]** |
| May-21 | **1.36**<br>**[1.28, 1.44]** | **1.34**<br>**[1.25, 1.42]** | **1.75**<br>**[1.63, 1.88]** | **1.68**<br>**[1.56, 1.80]** |

**Table 4:** Fully adjusted and unadjusted model results for depression and nervousness in Period 2 (N = 152,767)). The unadjusted models include one single covariate, selected from pandemic-related worries, demographics, and time. In contrast, the fully adjusted models include both the two worry variables and other covariates (demographics and time) together. Both model types integrate survey weights within logistic regression. Odds ratios are displayed as estimates with corresponding 95% Wald confidence intervals in the form of estimates [95% confidence interval], using a robust sandwich estimator for variance calculation. Significant results from Wald tests (significance level: 0.05) are highlighted in red and bold. Abbreviations: Ref, reference group; HS, high school.

|  | Depression |  | Nervousness |  |
| --- | --- | --- | --- | --- |
|  | Unadjusted | Adjusted | Unadjusted | Adjusted |
| **Gender**<br>(Ref: Male) |  |  |  |  |
| Female | **1.36**<br>[1.26, 1.48] | **1.50**<br>[1.37, 1.64] | **1.14**<br>[1.04, 1.26] | **1.21**<br>[1.08, 1.35] |
| **Age**<br>(Ref: 18-24) |  |  |  |  |
| 25-34 | **0.59**<br>[0.54, 0.64] | **0.64**<br>[0.58, 0.70] | **0.64**<br>[0.58, 0.71] | **0.70**<br>[0.62, 0.78] |
| 35-44 | **0.37**<br>[0.33, 0.41] | **0.42**<br>[0.38, 0.47] | **0.43**<br>[0.38, 0.48] | **0.49**<br>[0.43, 0.55] |
| 45-54 | **0.25**<br>[0.22, 0.29] | **0.31**<br>[0.27, 0.36] | **0.35**<br>[0.30, 0.40] | **0.42**<br>[0.36, 0.50] |
| 55-64 | **0.14**<br>[0.11, 0.18] | **0.20**<br>[0.16, 0.25] | **0.28**<br>[0.22, 0.35] | **0.38**<br>[0.30, 0.47] |
| 65+ | **0.14**<br>[0.09, 0.21] | **0.22**<br>[0.14, 0.34] | **0.20**<br>[0.15, 0.28] | **0.29**<br>[0.21, 0.40] |
| **Education**<br>(Ref: HS or More) |  |  |  |  |
| Less than HS | **1.60**<br>[1.44, 1.77] | **1.24**<br>[1.11, 1.39] | **1.50**<br>[1.34, 1.67] | **1.17**<br>[1.04, 1.32] |
| **Residential Status**<br>(Ref: Urban) |  |  |  |  |
| Rural | **1.15**<br>[1.07, 1.23] | **0.86**<br>[0.79, 0.93] | **1.22**<br>[1.12, 1.33] | 0.96<br>[0.86, 1.06] |
| **Region**<br>(Ref: South) |  |  |  |  |
| Central | **1.15**<br>[1.03, 1.27] | **0.88**<br>[0.79, 0.99] | 0.99<br>[0.87, 1.12] | **0.77**<br>[0.68, 0.88] |
| East | **1.46**<br>[1.32, 1.62] | **1.16**<br>[1.04, 1.29] | **1.25**<br>[1.12, 1.41] | 1.00<br>[0.89, 1.14] |
| North | 1.08<br>[0.97, 1.20] | 0.92<br>[0.82, 1.04] | **0.88**<br>[0.78, 1.00] | **0.75**<br>[0.66, 0.86] |

Table 4 continued from previous page
|  | Depression |  | Nervousness |  |
| --- | --- | --- | --- | --- |
| Northeast | 0.98<br>[0.83, 1.15] | **0.82**<br>**[0.70, 0.97]** | 0.99<br>[0.82, 1.19] | 0.86<br>[0.71, 1.03] |
| West | **0.87**<br>**[0.78, 0.98]** | **0.84**<br>**[0.74, 0.94]** | **0.81**<br>**[0.71, 0.93]** | **0.78**<br>**[0.68, 0.89]** |
| **Occupation**<br>**(Ref: Agriculture)** |  |  |  |  |
| Education | 0.97<br>[0.84, 1.11] | 1.10<br>[0.94, 1.29] | 0.91<br>[0.77, 1.07] | 1.12<br>[0.93, 1.34] |
| Construction | 0.96<br>[0.81, 1.14] | 1.13<br>[0.94, 1.35] | 0.88<br>[0.71, 1.08] | 1.00<br>[0.80, 1.25] |
| Health | 0.92<br>[0.79, 1.08] | 1.19<br>[1.00, 1.41] | 1.10<br>[0.90, 1.34] | **1.48**<br>**[1.19, 1.83]** |
| Others | 0.99<br>[0.90, 1.09] | **1.24**<br>**[1.10, 1.39]** | 0.94<br>[0.84, 1.06] | **1.18**<br>**[1.03, 1.36]** |
| Tourism | 1.13<br>[0.82, 1.55] | **1.53**<br>**[1.11, 2.10]** | 1.20<br>[0.81, 1.79] | **1.65**<br>**[1.09, 2.51]** |
| Transportation | 1.12<br>[0.91, 1.37] | 1.17<br>[0.94, 1.46] | 1.03<br>[0.81, 1.29] | 1.06<br>[0.82, 1.36] |
| **Financial Stress**<br>**(Ref: Not Worried)** |  |  |  |  |
| Worried | **4.16**<br>**[3.88, 4.46]** | **2.52**<br>**[2.30, 2.77]** | **3.42**<br>**[3.15, 3.71]** | **1.91**<br>**[1.70, 2.15]** |
| **Food Insecurity**<br>**(Ref: Not Worried)** |  |  |  |  |
| Worried | **5.07**<br>**[4.67, 5.50]** | **2.32**<br>**[2.07, 2.60]** | **4.81**<br>**[4.38, 5.28]** | **2.71**<br>**[2.37, 3.11]** |
| **Time**<br>**(Ref: May-21)** |  |  |  |  |
| Jun-21 | 0.97<br>[0.82, 1.15] | 0.99<br>[0.84, 1.18] | 0.96<br>[0.79, 1.17] | 0.98<br>[0.80, 1.20] |
| Jul-21 | 0.89<br>[0.76, 1.05] | 0.91<br>[0.76, 1.08] | 0.99<br>[0.82, 1.20] | 1.01<br>[0.83, 1.23] |
| Aug-21 | 0.94<br>[0.79, 1.11] | 1.00<br>[0.84, 1.20] | 0.88<br>[0.72, 1.07] | 0.93<br>[0.76, 1.14] |
| Sep-21 | 0.90<br>[0.76, 1.07] | 0.96<br>[0.80, 1.14] | 0.83<br>[0.67, 1.01] | 0.87<br>[0.71, 1.07] |
| Oct-21 | 0.89<br>[0.75, 1.05] | 0.97<br>[0.82, 1.16] | 0.95<br>[0.78, 1.16] | 1.03<br>[0.84, 1.26] |
| Nov-21 | 0.86<br>[0.72, 1.02] | 0.89<br>[0.75, 1.07] | 0.84<br>[0.69, 1.03] | 0.87<br>[0.71, 1.07] |
| Dec-21 | **0.74**<br>**[0.61, 0.89]** | **0.76**<br>**[0.63, 0.92]** | **0.70**<br>**[0.57, 0.88]** | **0.72**<br>**[0.58, 0.90]** |

Table 4 continued from previous page
|  | Depression |  | Nervousness |  |
| --- | --- | --- | --- | --- |
| Jan-22 | **0.84**<br>**[0.71, 1.00]** | 0.85<br>[0.71, 1.02] | 0.83<br>[0.68, 1.02] | 0.84<br>[0.68, 1.03] |
| Feb-22 | 0.97<br>[0.81, 1.17] | 1.03<br>[0.85, 1.25] | 0.99<br>[0.80, 1.22] | 1.03<br>[0.83, 1.29] |
| Mar-22 | 1.00<br>[0.83, 1.21] | 1.08<br>[0.89, 1.32] | 0.90<br>[0.72, 1.12] | 0.94<br>[0.75, 1.18] |
| Apr-22 | 0.93<br>[0.77, 1.12] | 1.01<br>[0.83, 1.22] | 0.85<br>[0.69, 1.06] | 0.90<br>[0.72, 1.12] |
| May-22 | 0.98<br>[0.80, 1.20] | 1.03<br>[0.83, 1.27] | 0.92<br>[0.72, 1.18] | 0.96<br>[0.74, 1.23] |
| Jun-22 | 1.01<br>[0.82, 1.24] | 1.14<br>[0.93, 1.41] | 1.02<br>[0.80, 1.30] | 1.12<br>[0.87, 1.44] |

In Period 2, the influence of demographic variables on depression and nervousness mirrored the results observed in Period 1, with significant associations identified for gender, age, residential status, geographic region, and occupation after adjusting for other factors (Table 4). This period also highlighted the significance of education, a factor not previously noted in Period 1. Notably, the fully adjusted models showed that individuals without a high school diploma had a 1.24 (95% CI: [1.11, 1.39]) and 1.27 (95% CI: [1.04, 1.32]) times higher odds of experiencing depression and nervousness, respectively, compared to individuals with at least a high school education.

### 3.4 Effect modification by gender on the associations between worry variables and mental health outcomes

Our analysis explored potential modification of the associations between three pandemic-related worries on depression and nervousness by self-reported gender, by adding an interaction term between gender and each of the worry variables separately into our fully adjusted survey-weighted model. Figure 2 and Table S2 illustrate the odds ratios for depression and nervousness across different worry variables within each gender, after adjusting for other worries, demographic factors, and calendar time. Throughout both periods, every pandemic-related worry variable demonstrated a significant association with mental health problems in both genders from the fully adjusted survey-weighted models. In Period 1, females had a significantly lower odds ratio of reporting depression and nervousness compared to males regarding the same concern (p_int_ *<* 0.01, where p_int_ denotes the p-value corresponding to the gender by worry interaction term in the fully adjusted model). Specifically, in terms of concerns about financial status, the odds ratios for feeling depressed and nervous were 2.03 (95% CI: [1.88, 2.19]) and 1.74 (95% CI: [1.58, 1.91]) for females, respectively, lower than the corresponding odds ratios for males, which were 2.59 (95% CI: [2.49, 2.70]) and 2.02 (95% CI: [1.92, 2.12]). Similarly, the odds ratios regarding food insecurity for depression and nervousness were 1.21 (95% CI: [1.07, 1.36]) and 1.32 (95% CI: [1.16, 1.51]) for females, respectively, again lower than those for males, who showed odds ratios of 1.56 (95% CI: [1.49, 1.64]) for depression and 1.67 (95% CI: [1.57, 1.77]) for nervousness. In Period 2, the smaller sample size resulted in wider confidence intervals, leading to no statistically significant gender differences in the odds ratios for the worry variables (for depression, p_int_ = 0.2 in finance stress and p_int_ = 0.4 in food insecurity; for nervousness, p_int_ = 0.3 in finance stress and p_int_ = 0.7 in food insecurity), despite varying point estimates.

**Figure 2:**
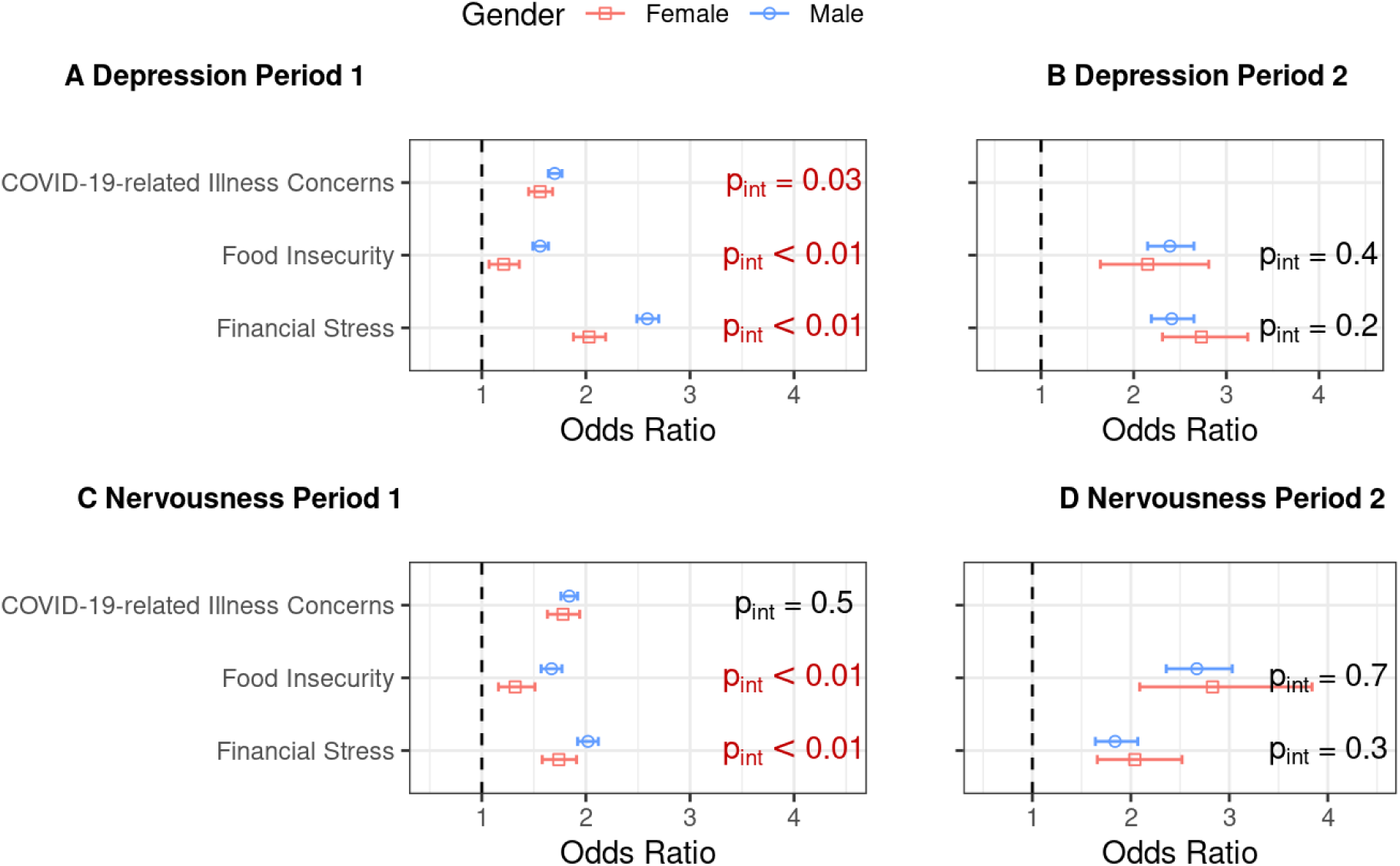
Effects of worry variables on mental health between genders in Period 1 (A: depression and C: nervousness; N = 595,229) and Period 2 (B: depression and D: nervousness; N = 152,767), post-weighting. The results were obtained from separate fully adjusted models that include a gender interaction term with one worry variable per model. Presented are odds ratios and their corresponding 95% Wald confidence intervals. Significant interaction terms (p_int_) from Wald tests (significance level: 0.05) are highlighted in red. All modes integrate survey weights within the logistic regression. Robust sandwich estimators were used for variance calculation. The vertical dashed line denotes the odds ratio of 1.

### 3.5 Effect modification by urban-rural residential status on the associations between worry Variables and mental health outcomes

We also examined how urban and rural residents were differently affected by three specific pandemic-related worries in terms of self-reported depression and nervousness. We added an interaction term for residential status and each worry variable separately into the fully adjusted model, accounting for survey weights. Figure 3 and Table S3 display the odds ratios for depression and nervousness associated with each worry variable within urban and rural settings, after controlling for other worries, demographic factors, and calendar time. Consistently, across both urban and rural contexts, each worry variable was significantly linked to mental health challenges. In Period 1, urban dwellers exhibited greater odds ratios of experiencing depression and nervousness regarding all of the three COVID-19-related worries than rural residents, with statistically significant interaction terms (p_int_ *<* 0.01) observed in all models. For instance, in terms of financial stress, urban individuals showed odds ratios of 2.44 (95% CI: [2.33, 2.56]) for depression and 1.98 (95% CI: [1.87, 2.09]) for nervousness, surpassing the odds ratios for rural residents, which stood at 2.12 (95% CI: [1.98, 2.26]) and 1.71 (95% CI: [1.58, 1.86]), respectively. In Period 2, urban residents continued to report significantly higher odds of depression and nervousness regarding these worries when compared to rural residents (for depression, p_int_ *<* 0.01 in finance stress; for nervousness, p_int_ *<* 0.01 in finance stress and p_int_ = 0.03 in food insecurity) expect the case with food insecurity for depression (p_int_ = 0.3). For example, in terms of worries about the financial situation, the odds ratios for urban individuals were 2.80 (95% CI: [2.52, 3.12]) for depression and 1.98 (95% CI: [1.73, 2.26]) for nervousness, which were higher than those for rural individuals, with odds ratios of 2.19 (95% CI: [1.92, 2.50]) for depression and 1.41 (95% CI: [1.20, 1.66]) for nervousness).

**Figure 3:**
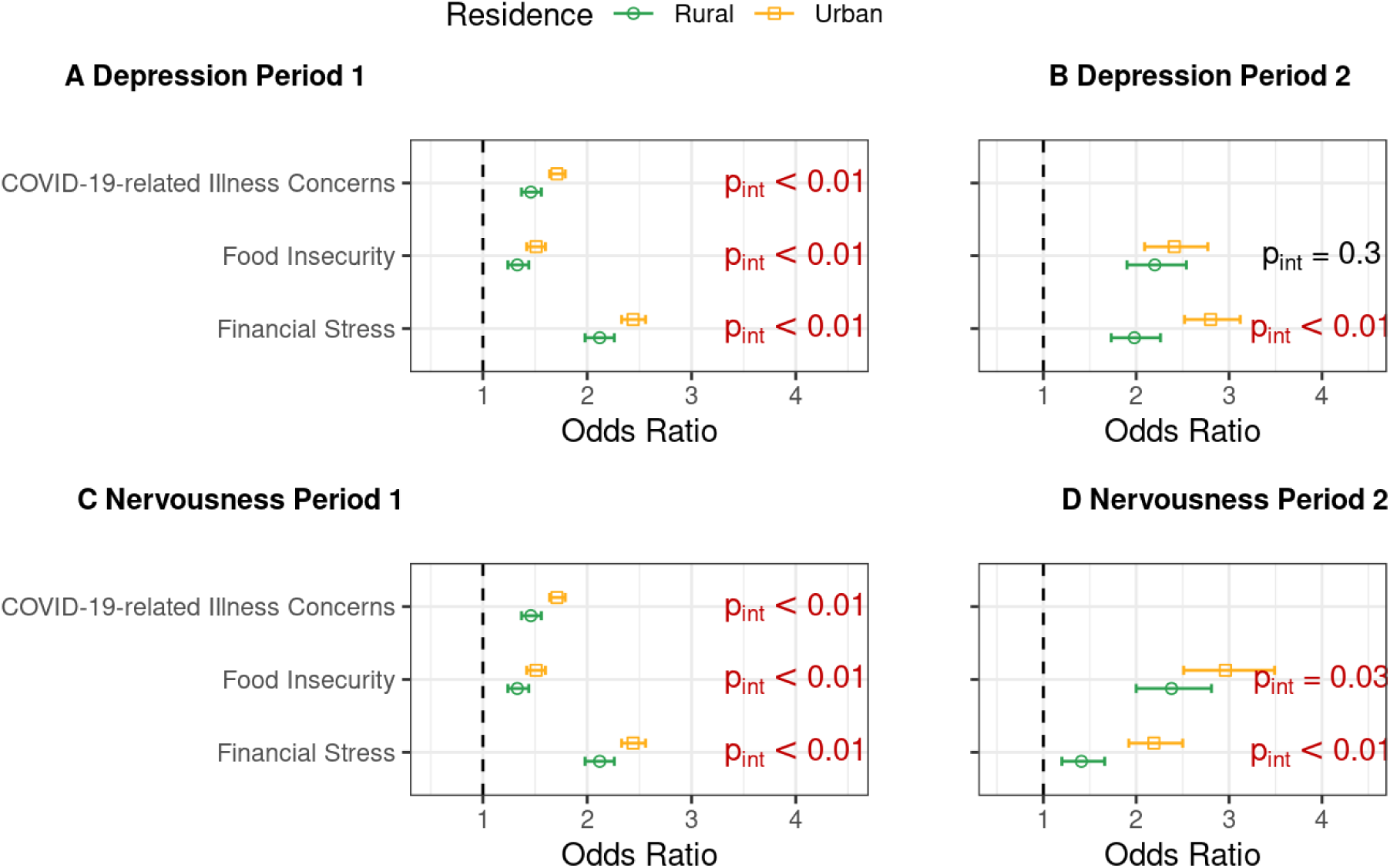
Effects of worry variables on mental health between urban and rural residential status in Period 1 (A: depression and C: nervousness; N = 595,229) and Period 2 (B: depression and D: nervousness; N = 152,767), post-weighting. The results were obtained from separate fully adjusted models that include a residential status interaction term with one worry variable per model. Presented are odds ratios and their corresponding 95% Wald confidence intervals. Significant interaction terms (p_int_) from Wald tests (significance level: 0.05) are highlighted in red. All modes integrate survey weights within the logistic regression. Robust sandwich estimators were used for variance calculation. The vertical dashed line denotes the odds ratio of 1.

### 3.6 Temporal variation in the associations between worry variables and mental health outcomes

We further delved into how the impact of the three subjective worries on depression and nervousness evolved over time by introducing interaction terms for time (categorized by month and year) with each worry variable individually into our fully adjusted survey-weighted models. Figure 4 presents the adjusted odds ratios for financial stress, food insecurity, and COVID-19-related concerns in relation to feeling depressed and nervous across time, in both periods. Vertical dashed lines represent the timing of the three peaks in newly confirmed COVID-19 cases and COVID-19-related deaths (September 2020, May 2021, and January 2022). During Period 1, financial stress emerged as the most influential factor affecting both depression (the range of odds ratios: [2.02, 2.71]) and nervousness (the range of odds ratios: [1.59, 2.19]), after adjusting for other variables. Although no discernible temporal patterns were evident for financial stress and food insecurity, there was a notable upswing in the odds ratios regarding COVID-19-related health concerns of reporting depression (odds ratio: 2.13, 95% CI: [1.90, 2.39]) and nervousness (odds ratio: 2.25, 95% CI: [1.98, 2.56]) during the second wave in May 2021 compared to the previous month (odds ratio 1.62, 95% CI: [1.46, 1.80] for depression and odds ratio: 1.73, 95% CI: [1.54, 1.94] for nervousness). During Period 2, the interpretation of time trends is constrained due to the smaller sample size, resulting in wider confidence intervals compared to Period 1. For nervousness, the point estimates for the effects of food insecurity (the range of adjusted odds ratios: [2.34, 3.75]) consistently surpassed those for financial stress (the range of adjusted odds ratios: [1.60, 2.43]) over time, while no significant differences were observed between these two sources of worry concerning depression.

**Figure 4:**
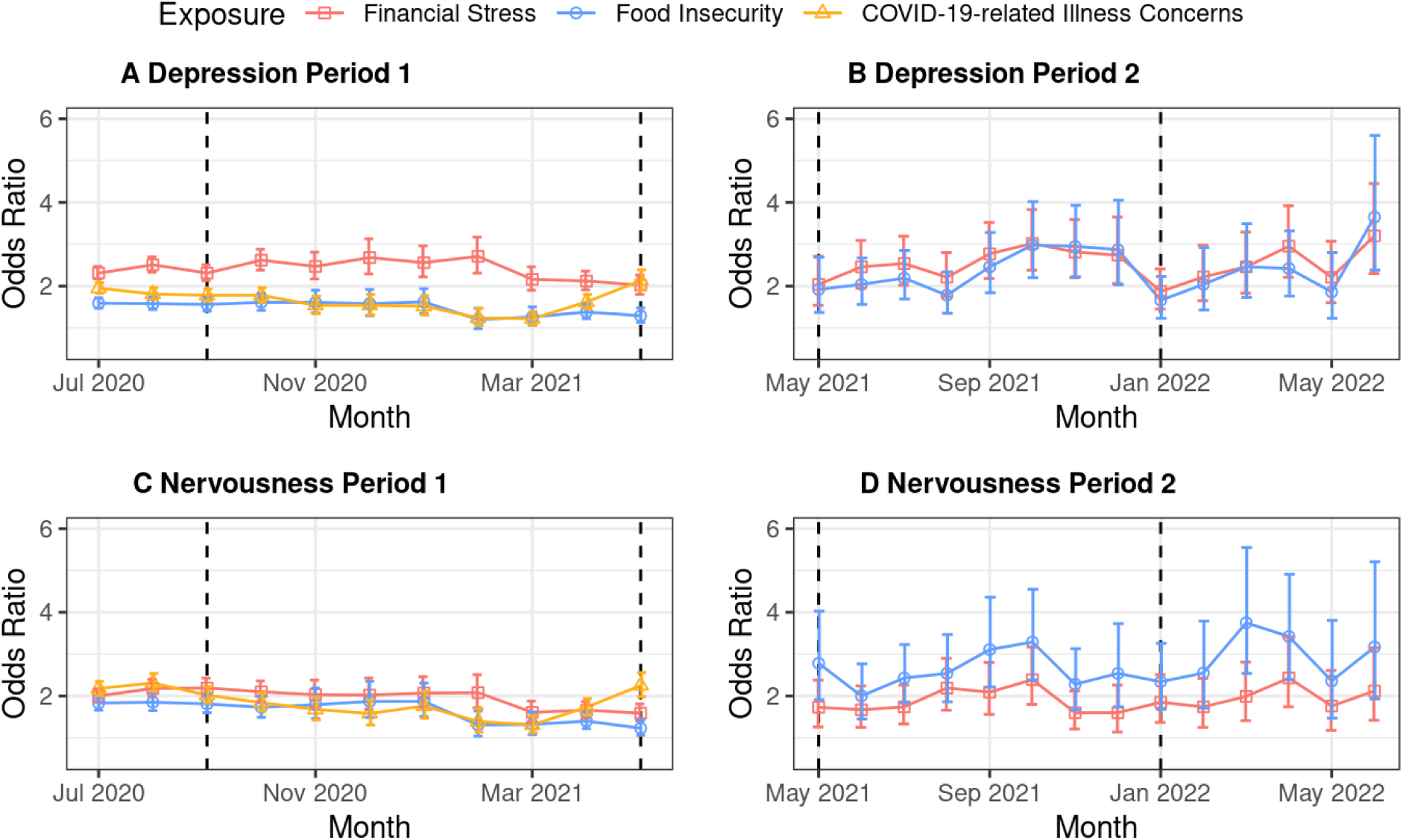
Effects of worry variables on mental health over time in Period 1 (A: depression and C: nervousness; N = 595,229) and Period 2 (B: depression and D: nervousness; N = 152,767), post-weighting. The results were obtained from separate fully adjusted models that include time interactions with one worry variable per model. Presented are odds ratios and their corresponding 95% Wald confidence intervals. All modes integrate survey weights within the logistic regression. Robust sandwich estimators were used for variance calculation. Vertical dashed lines denote the three peaks in new COVID-19 confirmed cases and related deaths in September 2020, May 2021, and January 2022, respectively. Due to limitations in sample size, data from June 2020 were merged with the July 2021 category.

## 4 Discussion

To our knowledge, this is the first study in India to examine mental health trends over an extensive period from 2020 to 2022, using a large-scale sample drawn from the nationwide general adult population. Our findings indicate a significant association between pandemic-related worries (financial stress, food insecurity, and COVID-19-related health worries) and symptoms of depression and nervousness. These results align with previous research conducted on specific subgroups within India (Kesar et al., 2021; Nguyen et al., 2021) and parallel findings from studies in other countries (Wilson et al., 2020a; Fang et al., 2021). Notably, financial stress emerged as the most significant factor affecting mental health during the first phase of our study period (June 27, 2020, to May 19, 2021) when adjusting for other worries, demographics, and time. Individuals concerned about their financial situation were found to have higher odds of experiencing depression (adjusted odds ratio: 2.36, 95% CI: [2.27, 2.46]) and nervousness (adjusted odds ratio: 1.91, 95% CI: [1.81, 2.01]), compared to those without such worries. This heightened impact of financial stress may be attributed to India’s economic structure, which is heavily dependent on low-wage informal labor and lacks robust social security measures to mitigate the negative effects of economic hardships among individuals (Ministry of Statistics and Programme Implementation, 2021). Our results advocate for the inclusion of financial allowances or subsidies and food rations within public health prevention strategies (Subbaraman et al., 2021). Effective policy-making should aim not only at saving lives from the disease but also at safeguarding livelihoods and ensuring social protection for the future.

Our findings underscore gender differences in the prevalence of mental health problems and their associations with pandemic-related worries. In the fully adjusted models but without interaction terms during Period 1, females had 1.52 (95% CI: [1.46, 1.59]) and 1.35 (95% CI: [1.28, 1.42]) times higher odds of depression and nervousness, respectively, than males. Introducing gender interaction terms in the fully adjusted models highlighted the effect modification by gender on the association between worry variables and self-reported mental health problems. Notably, during Period 1, females exhibited significantly lower odds ratios for both depression and nervousness when compared to males regarding the same pandemic-related worry. Such gender differences in odds ratios are consistent with findings from both COVID-19-related (Chatterji et al., 2021) and other research (Tran et al., 2018). These findings may reflect the influence of gender socialization roles (Fagot et al., 2012), where financial concerns might play a more central role in male social identities (Newcomb and Rabow, 1999). Additionally, females are often socialized to seek support more actively, which may serve as a protective factor in managing their worries (Denton et al., 2004). However, with respect to mental health outcomes, a large, multi-center study conducted by Kapoor et al. (2019) found marked gender discrimination in access to healthcare wherein men had 1.96 higher odds of visiting a tertiary healthcare center than women, which suggests comparatively fewer opportunities for mental health screening for women.

Our analysis revealed distinct urban-rural differences in the reported rates of depression and nervousness, along with how these mental health problems were linked to worry variables. Specifically, in Period 1 and without including interaction terms in our fully adjusted survey-weighted models, we observed that rural individuals were less likely to report depression and nervousness compared to urban individuals, with odds ratios of 0.82 (95% CI: [0.79, 0.85]) for depression and 0.89 (95% CI: [0.84, 0.93]) for nervousness. These findings align with similar observations made in China during the COVID-19 pandemic (Liu et al., 2021) and previous research conducted in India before the pandemic (Das et al., 2012; Ganguli, 2000). Furthermore, our results indicate that urban residents had higher odds ratios of self-reported mental health problems than rural residents regarding the same pandemic-related concern. This increased vulnerability among urban dwellers may stem from greater exposure to policies enforcing social isolation (Liu et al., 2021). COVID-19-related research in India has also underscored the significant impact of the pandemic on immigrant workers in urban areas, who were among the most affected groups (Kumar et al., 2020; Kesar et al., 2021) Additionally, we investigated three-way interactions involving gender, residential status, and worry variables (Table S4 and Table S5). However, the interpretability of these findings is constrained by the relatively small sample size available for each specific group.

We further examined changes over time in the prevalence of self-reported depression and nervousness and their association with pandemic-related worries. Notably, during the second wave of the pandemic in May 2021, there was a marked increase in the number of individuals experiencing depression or nervousness. The adjusted odds ratio for depression and nervousness regarding COVID-19-related health concerns also showed a substantial increase. This suggests that as the number of new cases and deaths sharply increased, people became more susceptible to mental health problems due to their heightened fears of infection. This period (May 2021) coincided with a severe public health crisis in India, marked by an overwhelmed healthcare infrastructure and a dramatic rise in hospital admissions (Kuppalli et al., 2021). The crisis was further exacerbated by the emergence of the B.1.617.2 sub-lineage, first identified in India, which the World Health Organization classified as a Variant of Concern in early May (World Health Organization, 2021).

In our analysis, we divided the study timeframe into two distinct periods, with the demarcation set for May 20, 2021, due to significant alterations in the format of the survey. The exploratory and statistical analysis revealed some discrepancies between these intervals. Specifically, the weighted prevalence rates for depression and nervousness in 2022 were substantially higher (8.29% for depression and 6.17% for nervousness) compared to those in 2020 (7.24% for depression and 4.77% for nervousness). Additionally, we observed variations in the odds ratio point estimates for worry variables. It is crucial to acknowledge the different phases of the pandemic these periods represent. For instance, the general availability of the COVID-19 vaccine in India commenced on May 1, 2021 (CNN, 2021), coinciding with the end of Period 1. Existing literature highlights the impact of COVID-19 vaccination on mental health outcomes (Perez-Arce et al., 2021; Rubinstein et al., 2023), suggesting varying vaccination rates may contribute to the differences observed between the periods. Furthermore, external factors unrelated to the pandemic, such as political events like a general election, may also affect the prevalence of mental health disorders (Krishna and Uvais, 2023). Therefore, careful consideration is necessary when interpreting and comparing the findings across these distinct periods.

Our study has a notable limitation due to its reliance on a non-probability, self-selected group of participants. Given the large sample size, the Big Data Paradox suggests that our prevalence estimates could significantly deviate from the true population values with a narrow confidence interval (Meng, 2018). The weights provided by Facebook were insufficient to ensure the representativeness of the sample, and they only accounted for age and gender. As a matter of fact, the observed prevalence rates for depression (8%) and nervousness (6%) may not accurately reflect the estimands in the general population, as they fall significantly below estimates from previous studies (Hossain et al., 2021). Nonetheless, it is important to note that the primary aim of our study was not to ascertain accurate prevalence rates, but rather to explore the relationships between various variables and their changes over time. As highlighted in Yang et al. (2023), the Big Data Paradox is less pronounced when the estimand shifts from the overall population averages to successive and subgroup differences in population means. Another limitation is that mental health outcomes available in the CTIS are self-reported and as such should not be interpreted as the presence of a clinical diagnosis and/or medical history of depression or anxiety disorders. Likewise, the self-reported nature of gender and residential status could also introduce measurement errors. Additionally, the same respondents were not followed over time in the CTIS data, which precluded the application of formal longitudinal methods. Although future research could benefit from using probabilistic survey samples for greater representativeness of the general population with more rigorous screening, we argue that our study offers a detailed and informative insight into the trends in mental health throughout the pandemic period in India.

## Data Availability

All data used in the study are accessed in adherence to a Data Use Agreement (DUA) with the University of Maryland.

## 5 Acknowledgements

We extend our heartfelt thanks to the Social Data Science Center at the University of Maryland for their invaluable role in conducting the global CTIS and granting us access to the individual-level data. We also thank Maxwell Salvatore for his help in visualizations.

## S1 Appendix

### S1.1 Supplementary tables

**Table S1:** Question phrasing and initial answers in the survey for Period 1 and Period 2.

|  |  | Period 1 | Period 2 |
| --- | --- | --- | --- |
|  |  | Jun 2020 - May 2021 | Jun 2020 - May 2021 |
| Mental Health |  |  |  |
| Depression | Question | During the last 7 days, how often did you feel so depressed that nothing could cheer you up? |  |
|  | Responses | 1 = All the time<br>2 = Most of the time<br>3 = Some of the time<br>4 = A little of the time<br>5 = None of the time |  |
| Nervousness | Question | During the past 7 days, how often did you feel so nervous that nothing could calm you down? |  |
|  | Responses | 1 = All the time<br>2 = Most of the time<br>3 = Some of the time<br>4 = A little of the time<br>5 = None of the time |  |
| Worries About The Pandemic |  |  |  |
| Finance Stress | Question | How worried are you about your household's finances in the next month? |  |
|  | Responses | 1 = Very worried<br>2 = Somewhat worried<br>3 = Not too worried<br>4 = Not worried at al |  |
| Food Insecurity | Question | How worried are you about having enough to eat in the next week? |  |
|  | Responses | 1 = Very worried<br>2 = Somewhat worried<br>3 = Not too worried<br>4 = Not worried at al |  |
| COVID-19-related Illness Concern | Question | How worried are you that you or someone in your immediate family might become seriously ill from coronavirus (COVID-19)? |  |

Table S1 continued from previous page
|  |  | Period 1 | Period 2 |
| --- | --- | --- | --- |
|  | Responses | 1 = Very worried<br>2 = Somewhat worried<br>3 = Not too worried<br>4 = Not worried at al |  |
| **Demographics** |  |  |  |
| **Gender** | **Question** | What is your gender? |  |
|  | **Responses** | 1 = Male<br>2 = Female<br>3 = Other<br>4 = Prefer not to answer |  |
| **Age** | **Question** | What is your age? |  |
|  | **Responses** | 1 = 18-24 years<br>2 = 25-34 years<br>3 = 35-44 years<br>4 = 45-54 years<br>5 = 55-64 years<br>6 = 65-74 years<br>7 = 75 years or older |  |
| **Education** | **Question** | How many years of education have you completed? | What is the highest level of education that you have completed? |
|  | **Responses** | Open response:<br>number validation | 1 = No formal schooling<br>2 = Less than primary school<br>3 = Primary school completed<br>4 = Secondary school completed<br>5 = High school (or equivalent) completed<br>6 = College/ pre-university/ University completed<br>7 = University post-graduate degree completed |
| **Residential Status** | **Question** | How many years of education have you completed? |  |
|  | **Responses** | 1 = City<br>2 = Town<br>3 = Village or rural area |  |
| **Region** | **Question** | What is the administrative region where you are currently staying? |  |
|  | **Responses** | Categorical response:<br>states and union territories of India |  |

Table S1 continued from previous page
|  |  | Period 1 | Period 2 |
| --- | --- | --- | --- |
| Occupation | Question | What is the main activity of the business or organization in which you work? |  |
|  | Responses | 1 = Agriculture<br>2 = Buying and selling<br>3 = Construction<br>4 = Education<br>5 = Electricity/water/gas/waste<br>6 = Financial/insurance/real estate services<br>7 = Health<br>8 = Manufacturing<br>9 = Mining<br>10 = Personal services<br>11 = Professional/scientific/technical activities<br>12 = Public administration<br>13 = Tourism<br>14 = Transportation<br>15 = Other |  |

**Table S2:** Effects of worry variables on mental health between genders in Period 1 (N = 595,229) and Period 2 (N = 152,767), post-weighting. The results were obtained from separate fully adjusted models that include a gender interaction term with one worry variable per model. All model types integrate survey weights within logistic regression. Odds ratios are displayed as estimates with corresponding 95% Wald confidence intervals in the form of estimates [95% confidence interval], using a robust sandwich estimator for variance calculation. Significant odds ratios from Wald tests (significance level: 0.05) are highlighted in red and bold. Significant interaction terms (p_int_) from Wald tests (significance level: 0.05) are also highlighted in red and bold.

| Period 1 (N = 595,229) |  |  |  |  |  |  |
| --- | --- | --- | --- | --- | --- | --- |
|  | Depression |  |  | Nervousness |  |  |
| | Male | Female | $p_{\text{int}}$ | Male | Female | $p_{\text{int}}$ |
| Finance Stress | **2.59**<br>[2.49, 2.70] | **2.03**<br>[1.88, 2.19] | **<0.01** | **2.02**<br>[1.92, 2.12] | **1.74**<br>[1.58, 1.91] | **<0.01** |
| Food Insecurity | **1.56**<br>[1.49, 1.64] | **1.21**<br>[1.07, 1.36] | **<0.01** | **1.67**<br>[1.57, 1.77] | **1.32**<br>[1.16, 1.51] | **<0.01** |
| COVID-19-related Illness Concern | **1.70**<br>[1.64, 1.77] | **1.56**<br>[1.45, 1.68] | **0.03** | **1.84**<br>[1.76, 1.92] | **1.78**<br>[1.63, 1.94] | **<0.01** |
| Period 2 (N = 152,767) |  |  |  |  |  |  |
|  | Depression |  |  | Nervousness |  |  |
| | Male | Female | $p_{\text{int}}$ | Male | Female | $p_{\text{int}}$ |
| Finance Stress | **2.41**<br>[2.19, 2.65] | **2.73**<br>[2.31, 3.13] | 0.17 | **1.84**<br>[1.64, 2.07] | **2.04**<br>[1.66, 2.52] | 0.34 |
| Food Insecurity | **2.39**<br>[2.15, 2.65] | **2.15**<br>[1.64, 2.81] | 0.43 | **2.67**<br>[2.36, 3.03] | **2.83**<br>[2.09, 3.84] | 0.70 |

**Table S3:**
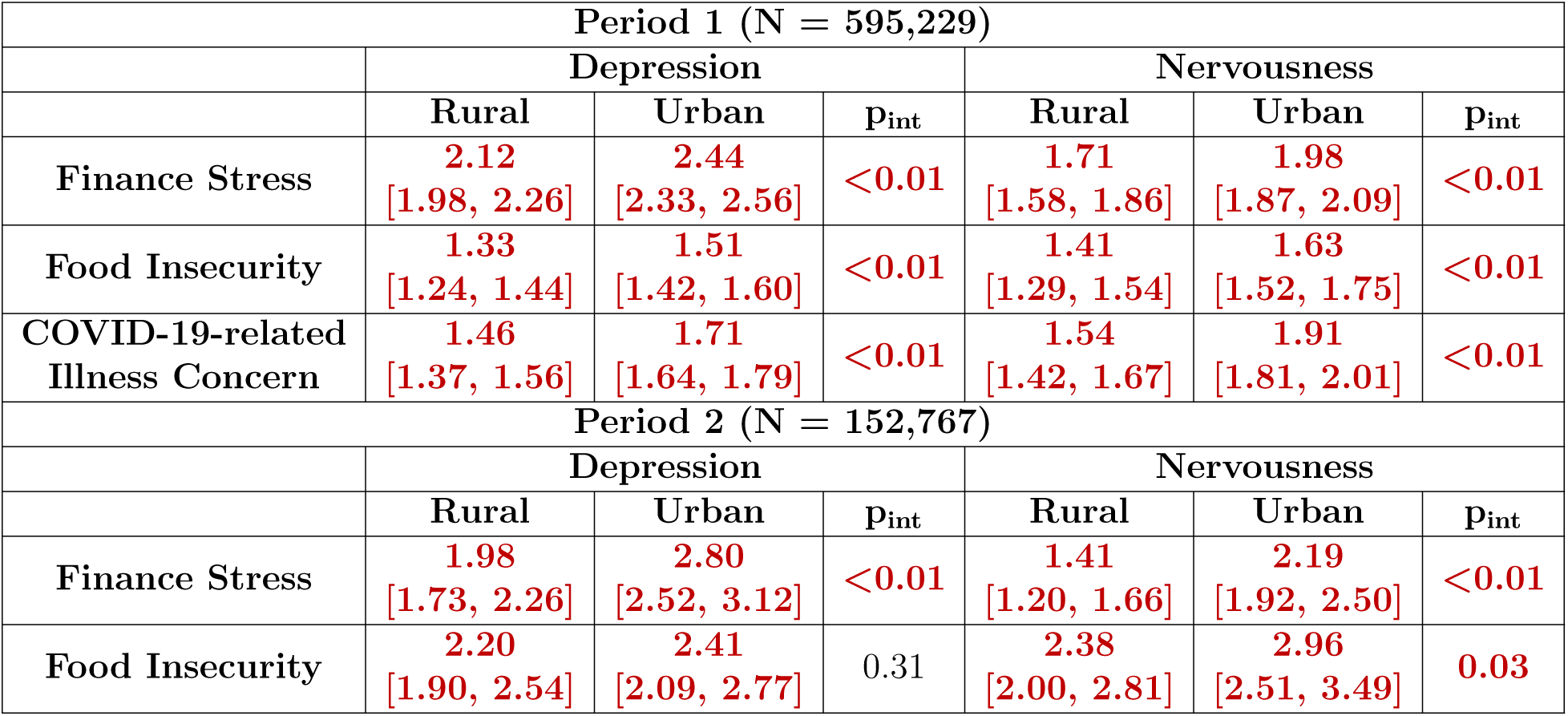
Effects of worry variables on mental health between urban and rural residential status in Period 1 (N = 595,229) and Period 2 (N = 152,767), post-weighting. The results were obtained from separate fully adjusted models that include a residential status interaction term with one worry variable per model. All model types integrate survey weights within logistic regression. Odds ratios are displayed as estimates with corresponding 95% Wald confidence intervals in the form of estimates [95% confidence interval], using a robust sandwich estimator for variance calculation. Significant odds ratios from Wald tests (significance level: 0.05) are highlighted in red and bold. Significant interaction terms (p_int_) from Wald tests (significance level: 0.05) are also highlighted in red and bold.

| Period 1 (N = 595,229) |  |  |  |  |  |  |
| --- | --- | --- | --- | --- | --- | --- |
|  | Depression |  |  | Nervousness |  |  |
| | Rural | Urban | $p_{int}$ | Rural | Urban | $p_{int}$ |
| Finance Stress | **2.12**<br>[1.98, 2.26] | **2.44**<br>[2.33, 2.56] | **<0.01** | **1.71**<br>[1.58, 1.86] | **1.98**<br>[1.87, 2.09] | **<0.01** |
| Food Insecurity | **1.33**<br>[1.24, 1.44] | **1.51**<br>[1.42, 1.60] | **<0.01** | **1.41**<br>[1.29, 1.54] | **1.63**<br>[1.52, 1.75] | **<0.01** |
| COVID-19-related Illness Concern | **1.46**<br>[1.37, 1.56] | **1.71**<br>[1.64, 1.79] | **<0.01** | **1.54**<br>[1.42, 1.67] | **1.91**<br>[1.81, 2.01] | **<0.01** |
| Period 2 (N = 152,767) |  |  |  |  |  |  |
|  | Depression |  |  | Nervousness |  |  |
| | Rural | Urban | $p_{int}$ | Rural | Urban | $p_{int}$ |
| Finance Stress | **1.98**<br>[1.73, 2.26] | **2.80**<br>[2.52, 3.12] | **<0.01** | **1.41**<br>[1.20, 1.66] | **2.19**<br>[1.92, 2.50] | **<0.01** |
| Food Insecurity | **2.20**<br>[1.90, 2.54] | **2.41**<br>[2.09, 2.77] | 0.31 | **2.38**<br>[2.00, 2.81] | **2.96**<br>[2.51, 3.49] | **0.03** |

**Table S4:** Effects of worry variables on mental health between genders among rural residents in Period 1 (N = 155,862) and Period 2 (N = 47,226), post-weighting. The results were obtained from separate fully adjusted models that include a gender interaction term with one worry variable per model. All model types integrate survey weights within logistic regression. Odds ratios are displayed as estimates with corresponding 95% Wald confidence intervals in the form of estimates [95% confidence interval], using a robust sandwich estimator for variance calculation. Significant odds ratios from Wald tests (significance level: 0.05) are highlighted in red and bold. Significant interaction terms (p_int_) from Wald tests (significance level: 0.05) are also highlighted in red and bold.

| Period 1 (N = 155,862) |  |  |  |  |  |  |
| --- | --- | --- | --- | --- | --- | --- |
|  | Depression |  |  | Nervousness |  |  |
| | Male | Female | $p_{int}$ | Male | Female | $p_{int}$ |
| Finance Stress | **2.26**<br>[2.10, 2.43] | **2.02**<br>[1.66, 2.46] | 0.28 | **1.87**<br>[1.71, 2.05] | **1.75**<br>[1.38, 2.22] | 0.59 |
| Food Insecurity | **1.49**<br>[1.37, 1.62] | 1.30<br>[0.99, 1.71] | 0.34 | **1.64**<br>[1.48, 1.82] | 1.32<br>[0.97, 1.81] | 0.18 |
| COVID-19-related Illness Concern | **1.57**<br>[1.46, 1.68] | **1.30**<br>[1.07, 1.58] | 0.07 | **1.69**<br>[1.55, 1.84] | **1.26**<br>[1.00, 1.60] | **0.02** |
| Period 2 (N = 47,226) |  |  |  |  |  |  |
|  | Depression |  |  | Nervousness |  |  |
| | Male | Female | $p_{int}$ | Male | Female | $p_{int}$ |
| Finance Stress | **1.72**<br>[1.48, 2.00] | **2.59**<br>[1.74, 3.83] | **0.05** | **1.34**<br>[1.10, 1.63] | 1.58<br>[0.94, 2.65] | 0.55 |
| Food Insecurity | **2.82**<br>[2.38, 3.33] | **2.92**<br>[1.78, 4.79] | 0.89 | **3.18**<br>[2.58, 3.91] | **3.48**<br>[1.87, 6.46] | 0.77 |

**Table S5:**
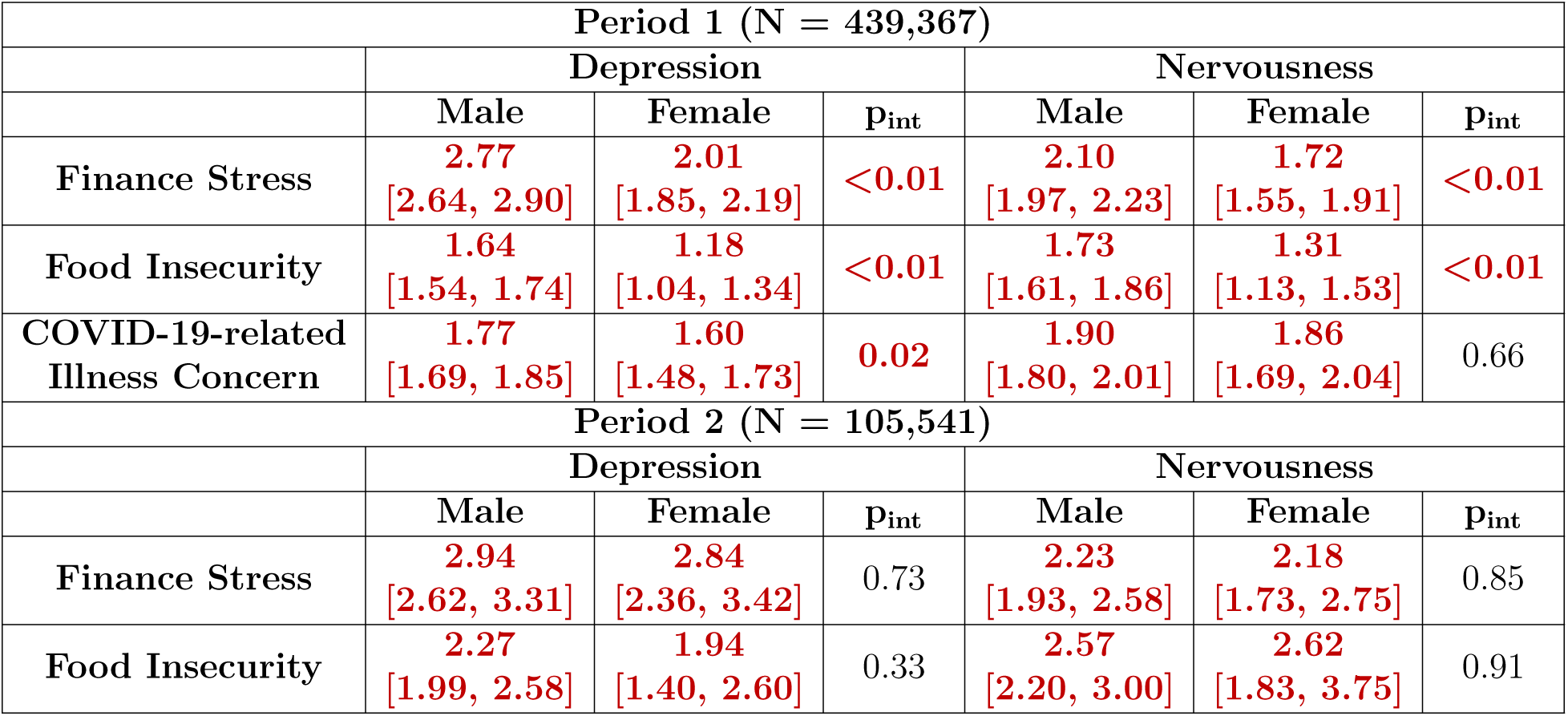
Effects of worry variables on mental health between genders among urban residents in Period 1 (N = 439,367) and Period 2 (N = 105,541), post-weighting. The results were obtained from separate fully adjusted models that include a gender interaction term with one worry variable per model. All model types integrate survey weights within logistic regression. Odds ratios are displayed as estimates with corresponding 95% Wald confidence intervals in the form of estimates [95% confidence interval], using a robust sandwich estimator for variance calculation. Significant odds ratios from Wald tests (significance level: 0.05) are highlighted in red and bold. Significant interaction terms (p_int_) from Wald tests (significance level: 0.05) are also highlighted in red and bold.

| Period 1 (N = 439,367) |  |  |  |  |  |  |
| --- | --- | --- | --- | --- | --- | --- |
|  | Depression |  |  | Nervousness |  |  |
| | Male | Female | $p_{int}$ | Male | Female | $p_{int}$ |
| Finance Stress | **2.77**<br>[2.64, 2.90] | **2.01**<br>[1.85, 2.19] | **<0.01** | **2.10**<br>[1.97, 2.23] | **1.72**<br>[1.55, 1.91] | **<0.01** |
| Food Insecurity | **1.64**<br>[1.54, 1.74] | **1.18**<br>[1.04, 1.34] | **<0.01** | **1.73**<br>[1.61, 1.86] | **1.31**<br>[1.13, 1.53] | **<0.01** |
| COVID-19-related Illness Concern | **1.77**<br>[1.69, 1.85] | **1.60**<br>[1.48, 1.73] | **0.02** | **1.90**<br>[1.80, 2.01] | **1.86**<br>[1.69, 2.04] | 0.66 |
| Period 2 (N = 105,541) |  |  |  |  |  |  |
|  | Depression |  |  | Nervousness |  |  |
| | Male | Female | $p_{int}$ | Male | Female | $p_{int}$ |
| Finance Stress | **2.94**<br>[2.62, 3.31] | **2.84**<br>[2.36, 3.42] | 0.73 | **2.23**<br>[1.93, 2.58] | **2.18**<br>[1.73, 2.75] | 0.85 |
| Food Insecurity | **2.27**<br>[1.99, 2.58] | **1.94**<br>[1.40, 2.60] | 0.33 | **2.57**<br>[2.20, 3.00] | **2.62**<br>[1.83, 3.75] | 0.91 |

### S1.2 Supplementary figures

**Figure S1:**
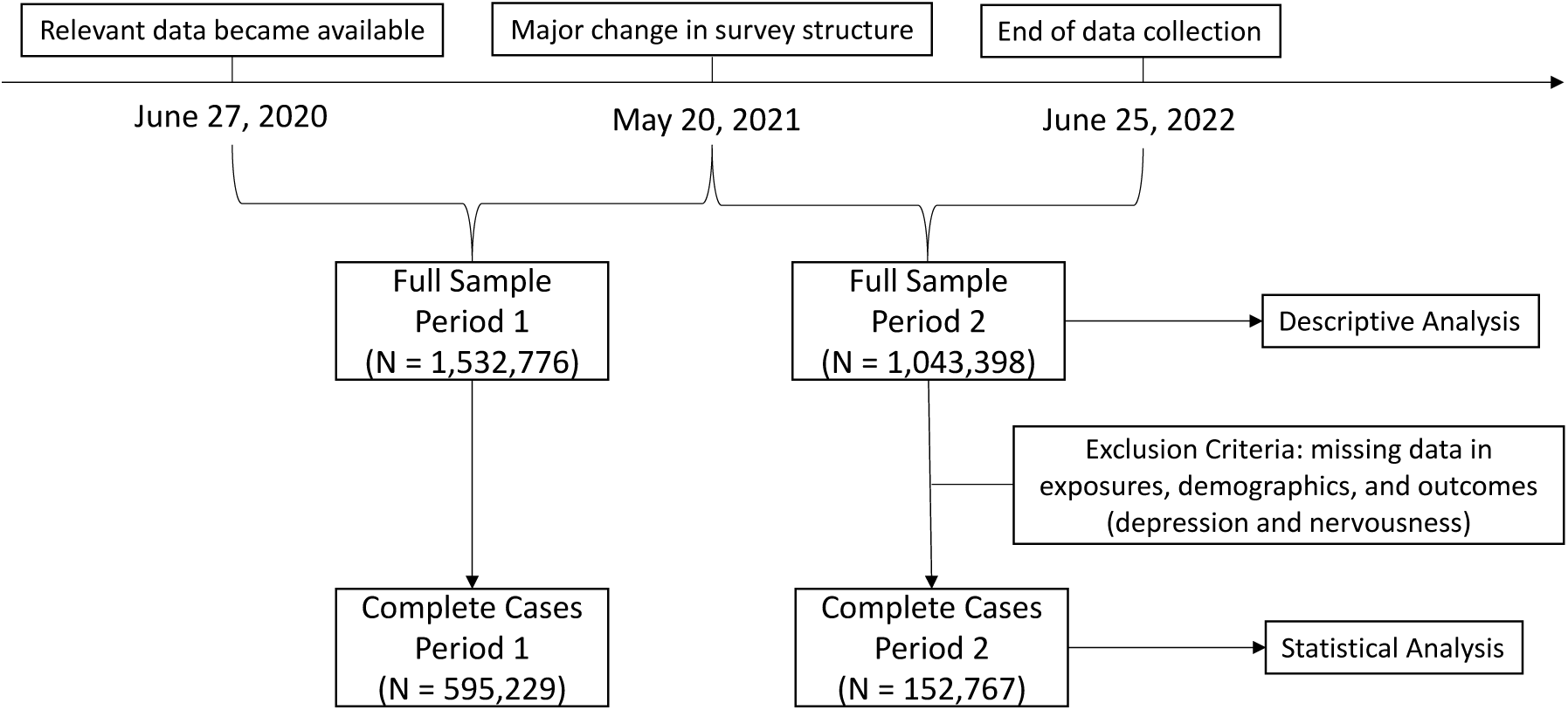
Overview of data processing. This study covers the timeframe from June 27, 2020, to June 25, 2022. A major change in the survey format on May 20, 2021, led to the division of the analytical period into two distinct phases. Descriptive analyses were conducted using the full dataset, whereas statistical evaluations were limited to cases with complete data.

